# Association between lockdown and changes in subspecies diversity with a focus on *Haemophilus influenzae* and *Staphylococcus aureus*

**DOI:** 10.1101/2024.05.08.24307047

**Authors:** Slim Hmidi, Nadim Cassir, Philippe Colson, Raymond Ruimy, Hervé Chaudet

## Abstract

**Background:** The social distancing measures implemented to curb SARS-CoV-2 transmission provided a unique opportunity to study the association between reduced human interaction and epidemiological changes related to human bacterial pathogens. While studies have indicated a decrease in respiratory infections during lockdowns, further description is needed regarding the changes in the incidence of bacterial populations. This study investigates the changes in strain richness of community infections with two bacterial species, *Haemophilus influenzae* and *Staphylococcus aureus* during the waning related to France’s social distancing measures, especially lockdown.

**Methods:** MALDI-TOF MS spectra analyses of routine clinical bacterial identifications were used as proxies for genomic analyses. Spectra from lockdown and reference periods were compared using unsupervised classification methods. A total of 251 main spectrum profiles of *H. influenzae*, 2079 main spectrum profiles of *S. aureus* for respiratory tract and blood samples, and 414 main spectrum profiles for skin samples of *S. aureus* were examined. Data were analyzed using hierarchical clustering, binary discriminant analysis, and statistical tests for significance.

**Results:** The strain mix of both bacteria during the lockdown was deeply altered, but with different further evolutions. *H. influenzae* exhibited a shift in spectra composition, with a subsequent return towards pre-lockdown diversity observed in 2021. In contrast, *S. aureus* exhibited a persistent change in spectra composition, with a gradual return to pre-lockdown patterns one year later.

**Conclusions:** Hindering inter-human transmission, as was done during the lockdown measures, was associated with significant alterations in bacterial species compositions, with differential impacts observed for *H. influenzae* and *S. aureus.* This study provides data on the putative relationship between genetic diversity and transmission dynamics during a public health crisis. Describing the dynamics of bacterial populations during lockdowns could contribute providing information for the implementation of future strategies for infectious disease control and surveillance.

## Introduction

To hinder the spread of the SARS-CoV-2 outbreak, several restrictive measures have been carried out worldwide, including population lockdowns to prevent viral transmission. In France, a general lockdown of the population was enforced from March 17^th^ to May 11^th^, 2020, for curbing the outbreak curve. This unprecedented situation can be considered as a real-scale experiment to assess the changes in the epidemiology of infectious agents when hindering human-to-human transmissions. This measure has been followed by less restrictive ones, as restrictions on gatherings and human mobility, use of personal protective equipment. However, the barrier gestures were less respected from summer 2020, allowing several epidemic waves.

Several studies have investigated the consequences of measures against the spread of SARS-CoV-2 on the incidence of other pathogens, mainly focusing on paediatric infections. Rotulo *et al.* demonstrated a reduction in community-acquired respiratory infections observed in a paediatric emergency department in Genoa, Italy, during the lockdown period 1. Similar results were also reported by Vierucci *et al.* in Tuscany 2.

A recent study examining invasive infections caused by *Streptococcus pneumoniae, Haemophilus influenzae* or *Neisseria meningitidis* across 26 countries during the early months of 2020 (from January 1 to May 31) retrieved a significant and prolonged decrease in invasive diseases due to these bacteria. These incidence reductions coincided with the implementation of COVID-19 containment measures in each respective region 3. Similarly, a study investigating the lockdown consequences on non-viral infectious agents observed significant changes in the relative abundance of several bacterial species, including a decrease in *Escherichia coli*, *S. pneumoniae*, and *H. influenzae* 4.

All these studies suggested a quantitative alteration of bacterial presence in the human population, with a decrease in species abundances. Interhuman transmission events have two dimensions: contact rate and inoculum size, constituting the transmission bottleneck. The lockdown, aimed at stopping or hindering the interhuman transmission, might result in small bottlenecks. Previous studies have shown that small bottlenecks were associated with substantially different levels of bacterial diversity in the transmission target, in terms of clonal richness, due to stochastic effects 5. Evaluating the epidemiological changes at the waning of the lockdown on human-related bacterial populations therefore need to assess species clonal richness over time.

Studies of bacteria subspecies optimally rely on costly genomic analyses. However, MALDI-TOF spectra analyses may act as proxies of genomic analyses. Routine identifications of bacterial species using MALDI-TOF (Matrix-Assisted Laser Desorption/Ionization Time-of-Flight) mass spectrometry are based on the presence of species’ characteristic peaks 6, a kind of ‘fingerprint recognition’, even if characteristic peak correspondence is not known 7. Identification may even delve deeper than the species level by the mean of supplementary peaks capable of identifying clonal complexes, frequently used for outbreak investigation and clonal characterization 8-11. We previously demonstrated 12 that MALDI-TOF spectra have sufficient richness and expressivity to allow for sub-species differentiations. On this basis, we have developed a set of original numerical and statistical processing tools for analyzing routine identification spectra to search for possible clonal complexes for epidemiologic surveillance purposes 13.

In this work we used these tools to address the following questions: what were the changes at the waning of the lockdown-related on species’ incidence and their clonal richness? Were these changes similar across bacterial species and samplings? This work specifically focused on *H. influenzae* and *Staphylococcus aureus*, species with enough identifications.

## Materials and methods

### Experimental design

The experimental design of this study aimed to compare MALDI-TOF spectra of *Haemophilus influenzae* and *Staphylococcus aureus* produced during the 8-week lockdown period with 8-week reference periods before and after, for identifying alterations in spectra clustering. This general framework was used to develop two protocols: the first protocol compared the lockdown period to the periods immediately before and following it within the same year, while the second protocol compared the lockdown period to the same one during 2019 and 2021.

### Materials

The Institut Hospitalo-Universitaire Méditerranée Infection (IHUMI) has a database of 1.270 million MALDI-TOF MS spectra carried out since 2011 for routine bacterial identification in the AP-HM, mostly from regional patient recruitment. All spectra were done using three Bruker Microflex mass spectrometers following the standardized protocol provided by Bruker Daltonics. A single colony or sediment was applied directly to two separate spots on crushed, air-dried steel targets coated with 1 μL of a matrix solution of α-cyano-4-hydroxycinnamic acid saturated in 50% acetonitrile and 2.5% trifluoroacetic acid, and air dried for 5 min. Spectrum acquisitions were controlled using FlexControl software. The Bruker Bacterial Test Standard (BTS) calibration was routinely used according to Bruker instructions. For each plate, BTS was used as a positive control and an uninoculated matrix solution was used as a negative control. Identification of bacterial species was given using the Biotyper software with species-level identification when the log (score) was ≥ 2.0. All MALDI-TOF spectra produced by the automated systems were loaded into a database, along with the corresponding biological sample information (pseudonymised patient demographics and stay characteristics, sample characterization, requesting unit, species identification, antibiogram).

### Inclusion criteria

We included spectra from routine clinical bacterial identification of *H. influenzae* and *S. aureus*, corresponding to upper respiratory tract and blood culture specimens for the two species and skin specimens for *S. aureus*. All samples were coming from community infections. We retained spectra with existing positive plate control and belonging to the 5 study periods: lockdown (March 17-May 11, 2020), pre-lockdown 2020 (January 27-March 16, 2020), post-lockdown (May 12-July 7, 2020), baseline 2019 (March 17-May 11, 2019), baseline 2021 (March 17-May 11, 2021). The spectra were deduplicated by hospital stay and clinical sample kind.

### MALDI-TOF MS analysis

The matrix gathering the intensities of the significant peaks was set up as recommended by S. Gibb 14. For spectra normalization we used a process flow made of intensity transformation (square root), smoothing (moving average with half window size 12), baseline correction (Statistics-sensitive Non-linear Iterative Peak-clipping algorithm, 100 iterations), and intensity recalibration on the peak of maximum intensity. Throughout this process, a signal-to-noise ratio (SNR) of 3 was utilized. The technical replicates were merged (averaging method) into main spectra profiles (msp), and species-specific peaks were removed to increase the contrast between spectra, as outlined in 12.

Our analysis was based on unsupervised classification, aiming at finding possible links between spectra compositions and their corresponding periods. Following the protocol we have described for epidemiological surveillance 13, we employed an ascendent hierarchical classification using Bray-Curtis distances between spectra, with Ward agglomeration and Gruveaus-Wainer ordination. The leaves of the dendrograms were color-coded to denote period membership (green: before the lockdown period, red: during, black: after, regardless of the protocol). During dendrogram analysis, the optimal number of clusters was determined using the Rousseeuw’s silhouette method 15, the homogeneity of period distribution among the clusters was tested using the chi2 or Fisher test, the grouping of lockdown strains was assessed using the Wald-Wolfowitz Runs Test (WWRT) 16, and we have systematically tested the proximity of lockdown and post-lockdown strains with Unconditional Barnard’s unilateral Exact Test (BET) on nearest neighbour counts, which is particularly suited to 2×2 table of counts 17. The most discriminating peaks between clusters were identified using the Gibb and Strimmer’s binary discriminant analysis 18.

The analysis was done using R and the packages MALDIquant v1.16.2, Seriation v1.2-2 for dendrogram ordering, BinDA v1.0.3 for binary discriminant analysis, and Vegan for distance metrics.

### Ethical approval statement

The surveillance system is in conformance with the Regulation (EU) 2016/679 of the European Parliament and of the Council of 27 April 2016 on the protection of natural persons with regard to the processing of personal data and on the free movement of such data, and repealing Directive 95/46/EC (General Data Protection Regulation), declared on the RGPD/AP-HM register with number 2019-73 (“Epidemiological monitoring of potentially pathogenic microorganisms”). The study SpectraSurv was approved by the *Commission Nationale Informatique et Libertés* (decision DR-2018-177). The data were accessed for research purposes during June 2022, and authors had no access to information that could identify individual participants during or after data collection.

## RESULTS

### Haemophilus Influenzae

#### Comparison with reference periods from the same year

We analysed 283 *H. influenzae* spectra grouped into 140 msp, and distributed as follows: 72 before lockdown (51.4% of all msp), 17 during lockdown (12.1%), 51 after lockdown (36.5%) (Fig 1a, colouring in accordance with the schema presented in the methods). According Rousseeuw’s methods, the dendrogram was partitioned into 5 clusters, heterogeneous in their composition regarding the periods, and contrasted by the presence of most of the lockdown samples in the lower subtree (Fisher test: p=0.0055). There was no grouping of lockdown strains together (WWRT=-1.96, p=0.051), but we observed a closer proximity of lockdown spectra with post-lockdown spectra (BET=-2.02, p=0.028)

**Fig 1:**
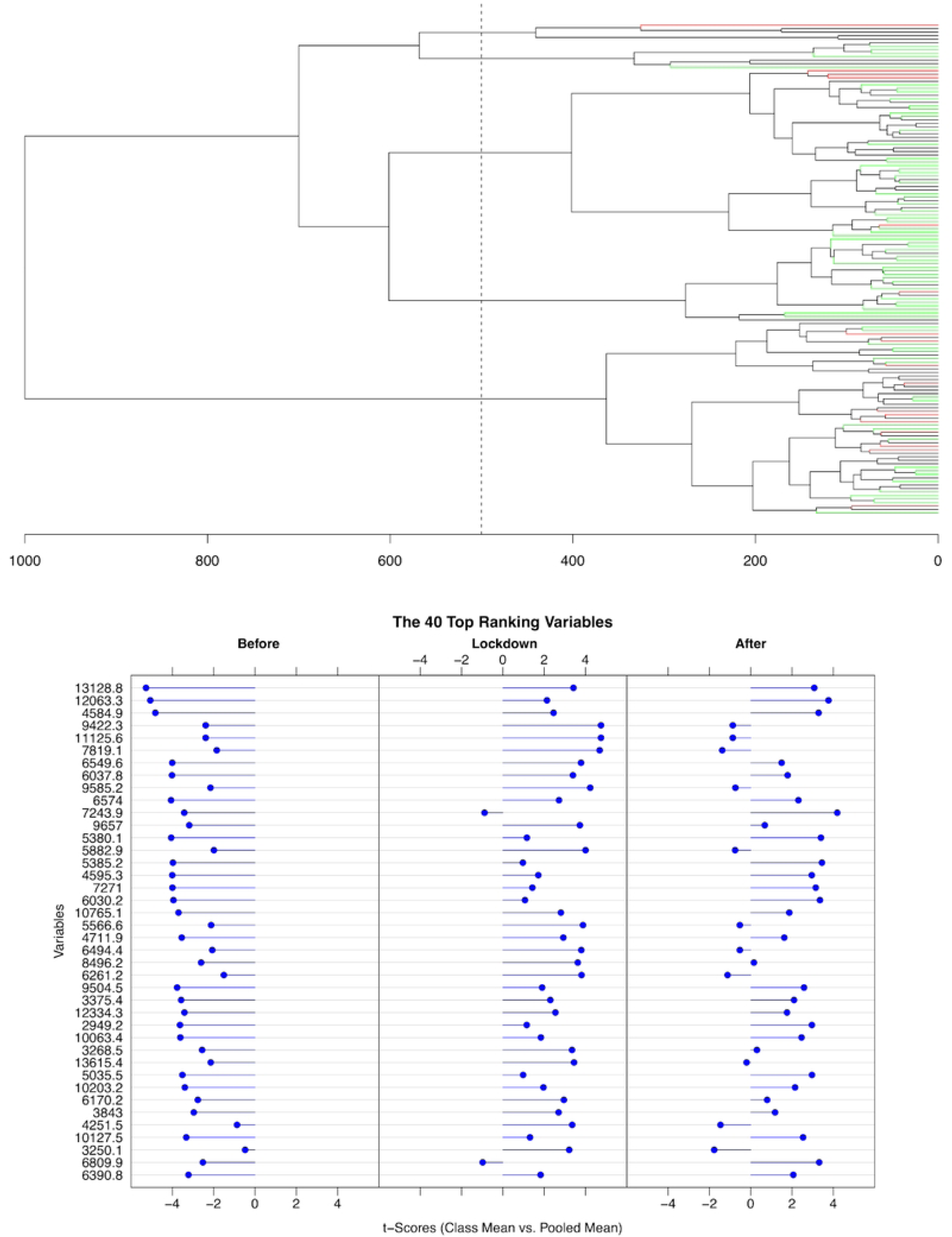
Results of the analysis of *H. influenzae* spectra from the same year. **a)** Dendrogram of the 140 main spectra profiles. The colour scale shows the distribution of the leaves over time: before lockdown (green), during lockdown (red) and after lockdown (black). The dotted line corresponds to the cut height for the optimal number of clusters. **b)** Binary discriminant analysis showing the 40 highest ranked peaks contrasting the lockdown period with the 8 weeks periods before and after lockdown. Peaks are indicated by their m/z. For each selected peak, the entropic t-score ranking is shown, positive when the peak is present in the group.

A binary discriminant analysis directly contrasting the spectra from the three periods showed an opposition of lockdown and pre-lockdown most discriminant peaks, with an important similarity of lockdown and post-lockdown periods (Fig 1b). This finding was in accordance with the proximity of spectra belonging to these two periods observed on the dendrogram and suggests that clonal complexes were replaced during the lockdown by new ones, persisting after the lockdown.

#### Comparison with years 2019 and 2021

We studied 260 spectra grouped into 128 msp, 73 (58.1%) from 2019, 19 (11.2%) from lockdown (2020), and 36 (30.7%) from 2021. The 3 clusters of the dendrogram (Fig 2a) were heterogeneous according to the period distributions with a single cluster gathering 13/19 lockdown strains (Chi2=12.015, p=0.017), and showed a grouping of the samples from the lockdown period (WWRT=-4.02, p=5.86 10^-5^). However, there was no privileged tying of lockdown strains with post-lockdown strains (BET=-1.37, p=0.097).

**Fig 2:**
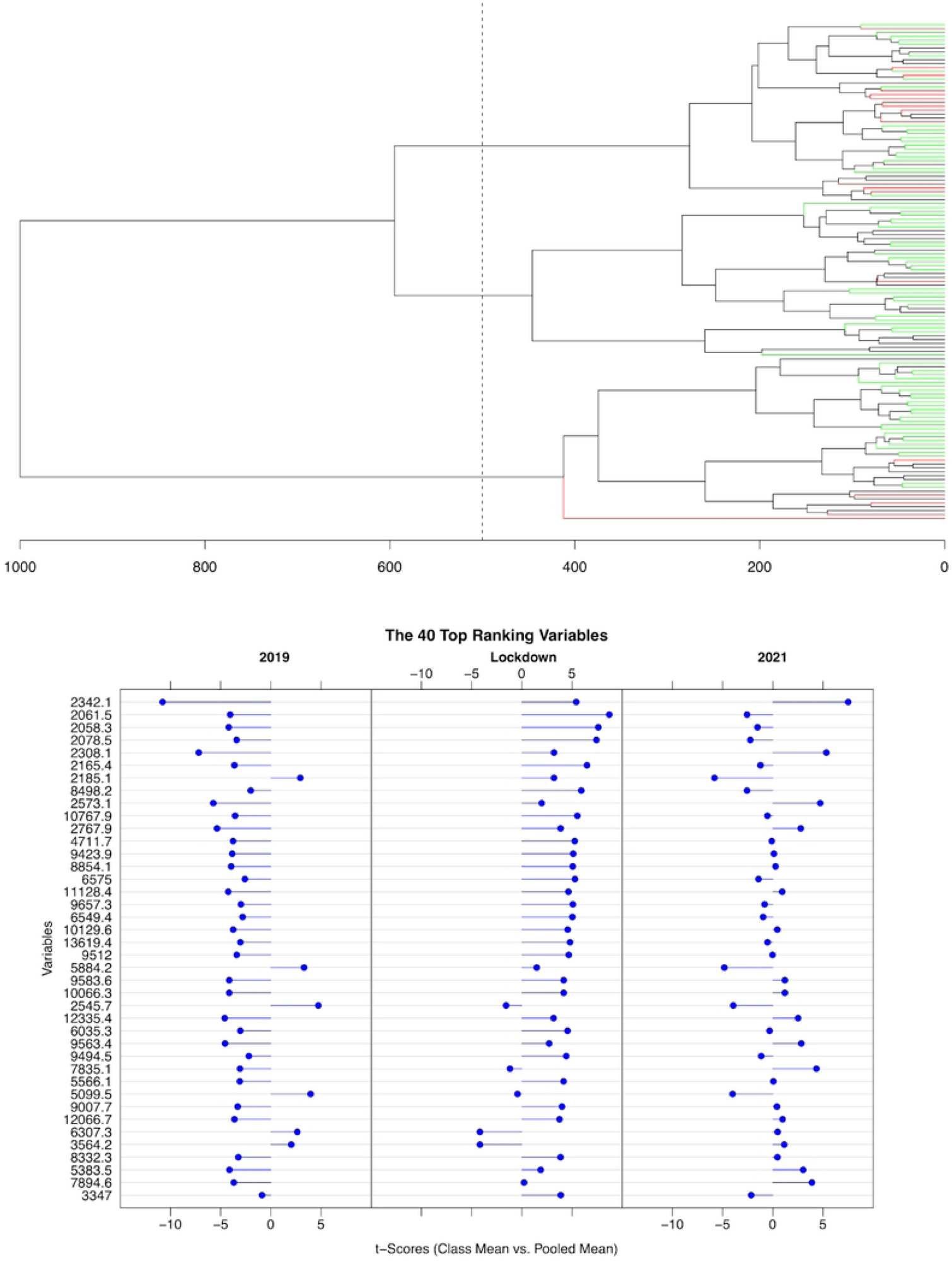
Results of the analysis of *H. influenza* spectra from the same period of the 3 years. **a)** Dendrogram of the 128 main spectra profiles. The colour scale shows the distribution of leaves over time: 2019 (green), 2020 (red) and 2021 (black). The dotted line corresponds to the cut height for the optimal number of clusters. **b)** Binary discriminant analysis showing the 40 highest ranked peaks contrasting the three yearly periods during 2019, 2020 (lockdown), and 2021. Peaks are indicated by their m/z. For each selected peak, the entropic t-score ranking is shown, positive when the peak is present in the group.

The binary discriminant analysis of the peaks contrasting these 3 periods (Fig 2b) showed that the most discriminating peaks appeared during the lockdown, but with a tendency during 2021 to return to the 2019 situation with an intermediate configuration. This result, associated with the absence of tying in the dendrogram, suggests that the clonal complex replacement observed during year 2020 was transient, with a slow and partial return to the pre-lockdown state. The two binary discriminant analyses shared 19/40 most discriminant peaks, showing a relative stability of the peaks supporting the discrimination.

### Staphylococcus aureus

For this species we also examined skin samples in addition to respiratory samples, due to the importance of the transmission mode by direct contact.

#### Respiratory tract and blood samples

##### Comparison with reference periods from the same year

We analysed 2147 spectra grouped into 1019 msp of *S. aureus* distributed as follows: 410 msp before lockdown represented in green (44.57% of the spectra), 200 msp during lockdown represented in red (18.37 %), 409 msp after lockdown represented in black (37.06%) (Fig 3a). Silhouette analysis of this dendrogram selected 6 clusters, heterogeneous according to the period distribution, with a single cluster having 32% of its strains belonging to the lockdown period and gathering 46% of these strains (Chi2=64.87, p=4.3 10^-10^). Lockdown strains were particularly grouped according to their spectra similarity (WWRT=-10.685, p<2.2 10^-16^). However, there was no privileged tying of lockdown strains with post-lockdown strains (BET=0.10, p=0.48).

**Fig 3:**
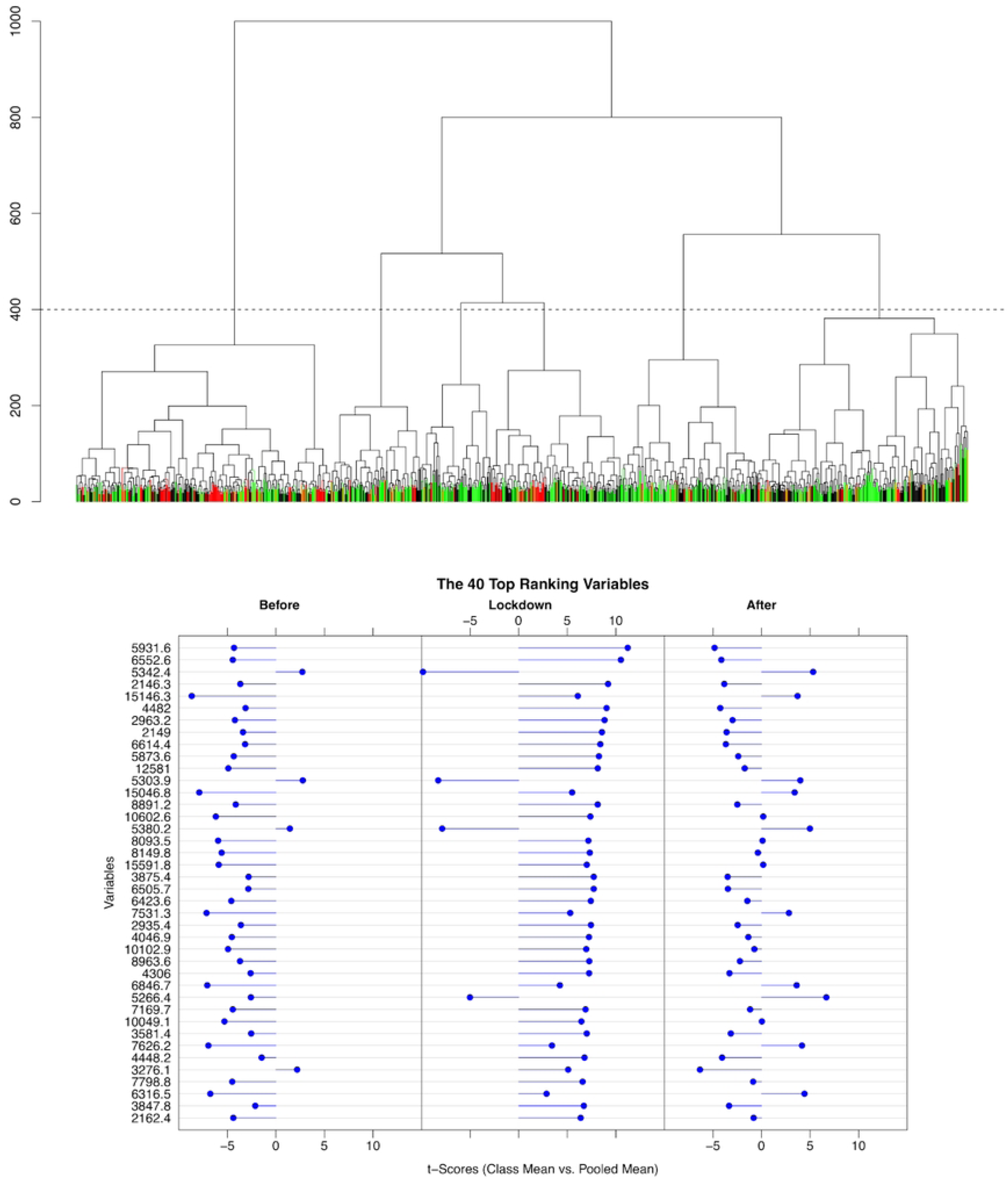
Results of the analysis of *S. aureus* spectra from respiratory tract and blood samples of the same year. **a)** Dendrogram of the 1019 main spectra profiles. The colour scale shows the distribution of the leaves over time: before lockdown (green), during lockdown (red) and after lockdown (black). The dotted line corresponds to the cut height for the optimal number of clusters. **b)** Binary discriminant analysis of main spectra profiles showing the 40 highest ranked peaks contrasting the lockdown period with the 8 weeks periods before and after lockdown. Peaks are indicated by their m/z. For each selected peak, the entropic t-score ranking is shown, positive when the peak is present in the group.

Like *H. influenzae*, the binary discriminant analysis comparing the spectra corresponding to the three periods (before, during, after lockdown) showed an opposition of the most discriminating peaks between the pre-lockdown and the lockdown period (Fig 3b). However, this opposition no more persisted during the post-lockdown period, with a partial evolution toward the pre-lockdown spectra characteristics.

##### Comparison with years 2019 and 2021

We analysed 2653 spectra grouped into 1260 msp of *S. aureus* distributed as follows: 475 (37.7%) msp from 2019, 200 (15.9%) msp from the lockdown period, 585 (46.4%) msp from 2021. Among the 3 dendrogram’s clusters confirmed by the silhouette analysis (Fig 4a), the middle one is characterised by a predominance of lockdown and post-lockdown strains at the expense of pre-lockdown ones (Chi2=79.801, p<2.2 10^-16^). As for the previous comparison, the lockdown strains were particularly grouped according to their spectra similarity (WWRT=-13.782, p<2.2 10^-16^), and we found a tying of lockdown and post-lockdown strains (BET=2.315, p=0.011).

**Fig 4:**
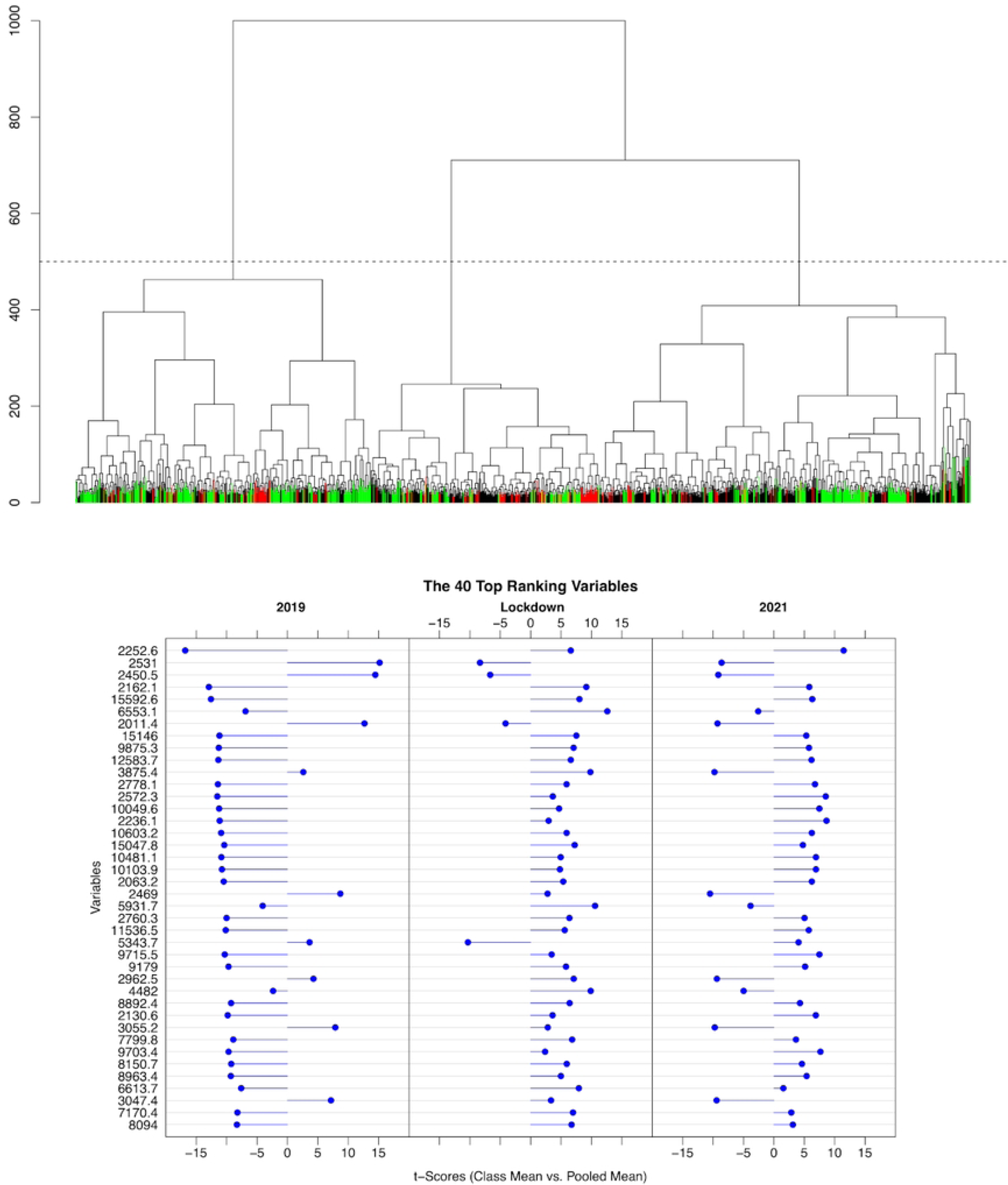
Results of the analysis of *S. aureus* spectra from respiratory tract and blood samples of the same period over the 3 years. **a)** Dendrogram of the 1260 main spectra profiles. The colour scale shows the distribution of leaves over time: 2019 (green), 2020 (red) and 2021 (black). The dotted line corresponds to the cut height for the optimal number of clusters. **b)** Binary discriminant analysis of main spectra profiles showing the 40 highest ranked peaks contrasting the three yearly periods during 2019, 2020 (lockdown), and 2021. Peaks are indicated by their m/z. For each selected peak, the entropic t-score ranking is shown, positive when the peak is present in the group.

The overall binary discriminant analysis (Fig 4b) showed a similarity between lockdown and 2021 and a difference from 2019, suggesting a persistence of the changes observed during the lockdown on the protein composition of *S. aureus*, which seems to agree with the results of the comparison between before and after it during the same year. The two binary discriminant analyses shared 21/40 most discriminant peaks, showing a stability of the peaks supporting the discrimination.

#### Skin Samples

##### Comparison with reference periods from the same year

This study analysed 367 msp including 177 msp before lockdown represented in green (48.2% of the spectra), 51 msp during lockdown represented in red (13.9 %), 139 msp after lockdown represented in black (37.9%). The silhouette analysis found three optimal clusters (Fig 5a) in the dendrogram, with a middle cluster having few lockdown strains (Chi2=10.949, p=0.027). Samples from the lockdown period were grouped by the clustering process of their spectra (WWRT=-8.506, p<2.2 10^-16^). We also noticed that spectra from pre-lockdown and post-lockdown samples were distributed over the dendrogram, without protein proximity between lockdown and post-lockdown periods (BT=-1.509, p=0.07).

**Fig 5:**
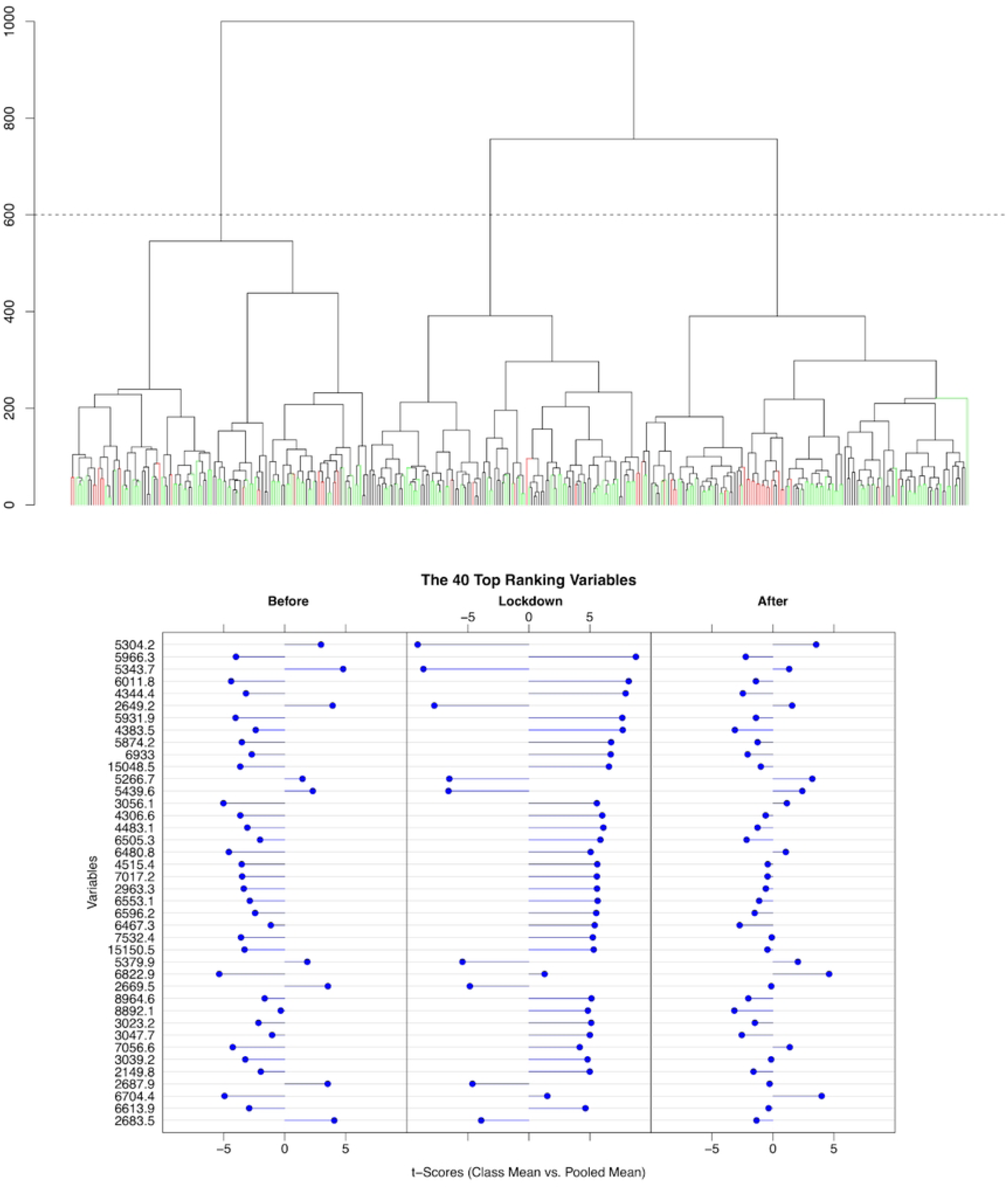
Results of the analysis of *S. aureus* spectra from skin samples of the same year. **a)** Dendrogram of the 367 main spectra profiles. The colour scale shows the distribution of the leaves over time: before lockdown (green), during lockdown (red) and after lockdown (black). The dotted line corresponds to the cut height for the optimal number of clusters. **b)** Binary discriminant analysis of main spectra profiles showing the 40 highest ranked peaks contrasting the lockdown period with the 8 weeks periods before and after lockdown. Peaks are indicated by their m/z. For each selected peak, the entropic t-score ranking is shown, positive when the peak is present in the group.

Unlike the previous comparison, the binary discriminant analysis (Fig 5b) showed a similarity of the distribution of the most discriminant peaks between pre and post lockdown spectra, contrasting with the lockdown period, which indicates that the changes during lockdown on the spectra composition of *S. aureus* were transitory.

##### Comparison with years 2019 and 2021

We analysed 605 spectra grouped into 288 msp of *S. aureus* distributed as follows: 221 (76.8%) msp from 2019, 51 (17.7%) msp from 2020, 16 (5.5%) msp from 2021. The two optimal clusters of the dendrogram (Fig 6a) presented a homogeneous distribution of the periods (Fisher Exact Test: p=0.656), but with a strong grouping of the lockdown strains (WWRT=-10.362, p<2.2 10^-16^) and a statistical association between lockdown and post-lockdown spectra despite the small amount of the last ones (BET=-1.715, p=0.048).

**Fig 6:**
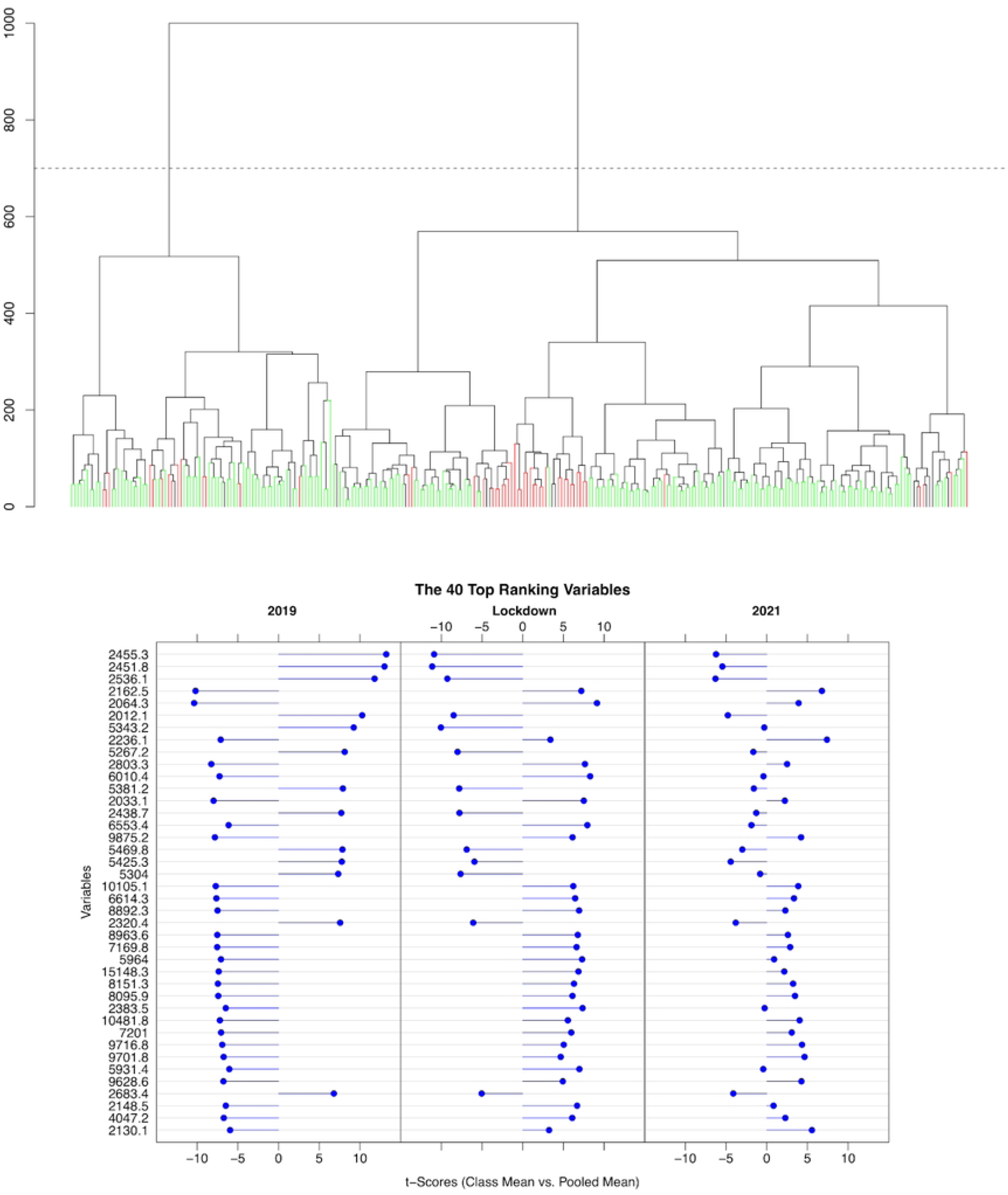
Results of the analysis of *S. aureus* spectra from skin samples of the same period over the 3 years. **a)** Dendrogram of the 1019 main spectra profiles. The colour scale shows the distribution of leaves over time: 2019 (green), 2020 (red) and 2021 (black). The dotted line corresponds to the cut height for the optimal number of clusters. **b)** Binary discriminant analysis of main spectra profiles showing the 40 highest ranked peaks contrasting the three yearly periods during 2019, 2020 (lockdown), and 2021. Peaks are indicated by their m/z. For each selected peak, the entropic t-score ranking is shown, positive when the peak is present in the group..

The overall binary analysis (Fig 6b) showed a similarity between lockdown and 2021 and a difference from 2019, suggesting a persistence of changes that occurred during the waning due to the lockdown on the protein composition of *S. aureus*, which seems to contradict the results of the comparison between before and after and during the same year, but agrees with the results of comparison with reference period from 2019 and 2021 for the respiratory tract and blood samples. The two binary discriminant analyses shared 14/40 most discriminant peaks, showing a change in the peaks supporting the discrimination.

## Discussion

Our objective was to evaluate the changes in bacterial species incidence during the waning due to the lockdown and associated social distancing measures, and their association with their clonal richness. This study focused on community strains of *Haemophilus influenzae* and *Staphylococcus aureus*, two species for which a notable and prolonged decrease was observed throughout this period. Bacterial identifications from blood and respiratory tract were pooled, as previous studies indicated that strains coming from these two kinds of samples could not be distinguished in most cases 19. Additionally, identifications of *S. aureus* from cutaneous samples were explored for comparison, as they are influenced by non-human transmissions 20. To our knowledge, no similar study, focusing on bacteria subspecies diversity, has been conducted thus far. For this purpose, we used MALDI-TOF MS, which is used for routine identification of bacterial species, and which is cost-effective compared to classic genotyping methods 21. This was possible because, in addition to species-specific peaks, MALDI-TOF MS spectra exhibit clusters of frequently co-occurring peaks, enabling subspecies differentiation 12.

This study included 251 msp of *H. Influenzae* and 2683 msp (2079 from respiratory sample, 604 from skin samples) of *S. aureus*, representing all identifications of these two species made during the included periods. All identifications were obtained from the routine work of the clinical microbiology laboratories of the four university hospitals of the Assistance Publique-Hôpital de Marseille. However, these results are limited to the AP-HM recruitment, which is primarily regional.

All six analyses of the two species revealed a dramatic change in the MALDI-TOF MS proteome composition during the lockdown, suggesting a shift in their strain composition. Regarding *H. influenzae*, this change persisted immediately after the lockdown, with a partial return in 2021 to a strain mix like the pre-lockdown situation. The frequent proximity in the dendrogram of strains coming from the lockdown and immediate post-lockdown periods suggests that subsequently encountered strains may have been selected during the lockdown, with a progressive increase in subspecies diversity over time. The grouping of lockdown spectra in the year comparisons, even if they are spread across several dendrogram clusters, supports the idea that they differ from the spectra of the other periods. Conversely, regardless of sample types, *S. aureus* displayed a change in spectral composition during the lockdown, but without a continuation during the immediate post-lockdown period. Similarities in spectra profiles with the lockdown period emerged later during the 2021 period, confirmed by statistically significant closer proximity of these spectra. While 21 out of 40 most discriminant peaks were shared by the two discriminant analyses of the respiratory and blood samples, indicating a resemblance of distinctive protein profile immediately around the lockdown period with the samples one year later, only 14 out of 40 most discriminant peaks were shared by the skin samples, showing a looser likeliness of spectral profiles one year later.

Interpreting the results requires consideration that the apparent population diversity conveyed by MALDI-TOF spectra reflects the species’ strain diversity, which is a trade-off between within-host diversities, mutations, inoculum strain heterogeneity, inoculum frequency, environmental selection, and transmission bottleneck. Previous simulation studies showed that small bottlenecks rapidly result in a single dominant strain carried and transmitted in the infected population 5. Our results indicated a decrease in strain diversity for both studied bacteria. An hypothesis is that the lockdown and preventive measures could have been associated with a reduction in inoculum frequency and an increase in the bottleneck effect due to reduced social interaction, and could have contributed to a reduction in genetic diversity within the bacterial population. Additionally, strict hygiene measures, such as frequent handwashing, use of disinfectants, and wearing masks, all distancing measures used during the end of 2020, were reported to reduce bacteria diversity 22. Household transmission could have become the primary mode of transmission, at the expense of community transmission.

*H. influenzae* is an obligate commensal of the human respiratory tract and non-typeable *H. influenzae* (NTHi) are currently the predominant strains 23. NTHi strains are more diverse and influenced by recombination compared to encapsulated *H. influenzae* 24. These genetic mechanisms, altering the gene expression or content, generate strains with fitness characteristics aimed at surviving under various selective pressure within a same kind of host 25. In particular, NTHi are able to switch between genetic variants at high frequency (phase variation mechanism) 26. This capability of dynamic recombination could explain why the change in *H. influenzae* strains’ diversity was only transitory for *H. influenzae*.

On the contrary, *S. aureus* is found in warm-blood animals and in the environment, constituting an important reservoir 20. It shows a slow but detectable genetic evolution of its population, at a rate of 2.72 mutations per megabase per year 19, in association with a fast replication rate 27. Twenty to twenty-five percent of the human population present a persistent carriage of *S. aureus* 28, which is rarely polyclonal (about 6.6 %) 29, mainly influenced by household transmission 20, possibly potentialized by host genetic factors 30. Epidemiologically isolated populations have shown very low diversity of their carriage 30. The rapid return to the pre-lockdown spectral configuration, followed by a progression toward the lockdown situation one year later, may be explained by a first influence of the reservoir multiplicity, followed by a return to the predominance of household transmissions, strengthen by the post-lockdown preventive measures.

This study showed that, for our human population and the studied bacteria species, lockdown was associated with a dramatic change of species compositions, but with different further evolution. We observed a transitory persistence of the strain composition that occurred during the lockdown for a human-dependant bacteria species, with a high frequency of genetic variant. Conversely, we observed a long-term evolution toward a return to the lockdown composition for a bacterial species with multiple reservoirs, a slow mutation rate, and an important household-dependant transmission.

## Data Availability

The data underlying the results presented in the study are available from the corresponding author

